# The mediating role of trust in physicians on the association between multidimensional health literacy and medication adherence in hemodialysis: A cross-sectional study

**DOI:** 10.1101/2023.09.04.23294984

**Authors:** Ryohei Inanaga, Tatsunori Toida, Tetsuro Aita, Yusuke Kanakubo, Mamiko Ukai, Takumi Toishi, Atsuro Kawaji, Masatoshi Matsunami, Tadao Okada, Yu Munakata, Tomo Suzuki, Noriaki Kurita

**Affiliations:** Department of Clinical Epidemiology, Graduate School of Medicine, Fukushima Medical University, Fukushima, Japan; Department of Nephrology, Shin-Yurigaoka General Hospital, Kanagawa, Japan; School of Pharmaceutical Sciences, Kyushu University of Health and Welfare, Miyazaki, Japan; Department of General Internal Medicine, Fukushima Medical University Hospital, Fukushima, Japan; Tessyoukai Kameda Family Clinic Tateyama, Chiba, Japan; Division of Clinical Epidemiology, Research Center for Medical Sciences, The Jikei University School of Medicine, Tokyo, Japan; Department of Nephrology, Kameda Medical Center, Chiba, Japan; Chikuseikai Munakata Clinic, Tokyo, Japan; Munakata Clinic, Chiba, Japan; Department of Innovative Research and Education for Clinicians and Trainees (DiRECT), Fukushima Medical University Hospital, Fukushima, Japan

**Keywords:** end-stage kidney disease, health literacy, medication adherence, trust in physicians

## Abstract

**Rationale & Objective:** Basic health literacy (HL) and trust in physicians can influence medication adherence (MA) in dialysis patients. However, how high-order HL is associated with MA, and how trust in physicians mediates this association remain unclear. We assessed the interrelationships between HL, trust in physicians, and MA, and investigated the mediating role of trust in physicians in the relationship between HL and MA.

**Study Design:** Multicenter cross-sectional study.

**Setting & Participants:** Japanese adults receiving outpatient hemodialysis at six dialysis centers.

**Exposures:** Multidimensional HL was measured using the 14-item Functional, Communicative, and Critical Health Literacy Scale. Trust in physicians was measured using the five-item Wake Forest Physician Trust Scale.

**Outcome:** MA was measured using the 12-item Adherence Starts Knowledge (ASK-12) scale.

**Analytical Approach:** A series of general linear models was created to analyze the associations between HL and ASK-12 scores with and without trust in physicians. Mediation analysis was performed to determine whether trust in physicians mediated this association.

**Results:** In total, 455 patients were analyzed. Higher functional and communicative HL were associated with lower barriers to MA (per 1-point increase: -1.90 (95% confidence interval (CI): -2.67, -1.13) and -2.11 (95% CI: -3.35, -0.87), respectively), whereas higher critical HL was associated with higher barriers (per 1-point increase: 1.67 (95% CI: 0.43, 2.90)). After controlling for trust in physicians, the magnitude of the association between HLs and MA decreased. Trust in physicians partially mediated the association between functional or communicative HL and MA (especially “beliefs”) and completely mediated the association between critical HL and MA (especially “behaviors”).

**Limitations:** Possible reverse causation.

**Conclusions:** In addition to functional HL, communicative and critical HL were associated with MA, and their associations were mediated by trust in physicians. To effectively improve MA, individualized strategies for each HL and favorable physician–patient interactions are important.

## Introduction

Adherence to routinely prescribed medications can pose significant challenges for patients with end-stage kidney disease (ESKD). These challenges arise because of the substantial number of medications involved and the difficulty in attributing specific effects and symptoms to individual drugs.^1^ Patients with ESKD often require 10–12 medications per day, consisting primarily of phosphorus adsorbents and antihypertensive drugs,^2,3^ and the average nonadherence rate to oral medication is reported to be 67%.^4^ The importance of medication nonadherence is highlighted by the inadequate control of mineral metabolism markers and increased risk of mortality.^5,6^ Health literacy (HL) among patients with ESKD may be an underlying cause of medication adherence (MA); studies have shown that HL plays an important role in self-care in patients with chronic kidney disease (CKD).^7^ However, there is insufficient research investigating which aspects of HL among patients with ESKD are connected to MA during their day-to-day interactions with healthcare providers.

In particular, communicative HL (referring to the ability to gather information through communication and apply new knowledge to changing situations) and critical HL (involving the ability to critically analyze information and use it to take control of a situation) are regarded as high-level HL based on functional HL, which is a basic skill in reading and writing that has received much attention.^8^ However, existing literature on the relationship between multidimensional HL and MA among patients with ESKD presents conflicting findings.^9,10^ For instance, a study involving a limited number of hemodialysis patients (<50 years) found associations between all three dimensions of HL and MA. However, the study had limitations in terms of the sample size and lack of validation of the MA assessment.^10^ In another study involving patients with CKD, including those with ESKD, MA was evaluated using a validated scale; however, no association was found between MA and the encompassing dimensions.^9^

Furthermore, there is a lack of research exploring how these dimensions of HL may influence physician–patient interactions and subsequently impact MA. This interaction is considered a key mediator between HL and self-care, such as MA,^11^ and trust in physicians plays a central role in such interactions.^12^ Indeed, trust in physicians has been shown to improve MA in patients with non-renal chronic diseases.^13^ However, although communicative HL has been found to foster trust in physicians for non-renal chronic diseases,^14^ studies on CKD have only generated hypotheses through one qualitative study.^15^ Examining whether and how these broad dimensions of HL can enhance MA through trust in physicians can help us to consider what level of HLs can be tailored to support individual patient needs and whether this support can be applied to promote MA within and beyond the context of physician–patient interactions.

Therefore, the present study aimed to accomplish two objectives within the context of Japanese hemodialysis patients: (1) explore the interrelationships between HL (functional, communicative, and critical HL), trust in physicians, and MA, and (2) investigate the mediating role of trust in physicians in the relationship between HL and MA using mediation analysis.

## Methods

### Study design and subjects

This cross-sectional multicenter study was conducted at six medical facilities providing outpatient hemodialysis services. This study adhered to the Declaration of Helsinki and was approved by the Ethical Review Board of Fukushima Medical University (number: ippan2021-292). The inclusion criteria were as follows: (1) adult patients with ESKD treated with hemodialysis or a hybrid treatment of hemodialysis and peritoneal dialysis; (2) regular visits to the participating institution for dialysis treatment; and (3) the ability to respond to the questionnaire. Patients (1) without regular medication prescriptions and (2) with severe dementia or complete blindness were excluded. Written consent was obtained from all participants. Paper-based questionnaires were completed by the participants and collected by medical staff without identifying the content. Respondents were offered the financial incentive of a 500-yen gift card. Data were collected between April 2022 and February 2023.

### Measures

#### MA

The main outcome was MA, measured using the Japanese version of the Adherence Starts with Knowledge (ASK)-12 scale.^16,17^ The ASK-12 is a 12-item questionnaire that measures MA in three domains: inconvenience/forgetfulness (three items), treatment beliefs (four items), and behavior (five items) (Box S1). Each item is scored on a five-point scale, with higher scores indicating greater adherence difficulties. Items 4–7 were scored in reverse to align with the other items. The total score is the sum of all items and ranges from 12 to 60. The validity and consistency reliability (coefficient alpha: 0.75) of the ASK-12 were confirmed,^16^ and the criterion validity of the Japanese version of the ASK-12 in relation to pharmacy refill rates and exacerbation frequency was demonstrated among patients with asthma.^17,18^

#### HL

The main exposure was measured using the Functional, Communicative, and Critical Health Literacy (FCCHL) scale, which is a self-reported measure that separates functional, communicative, and critical HL components.^19^ The scale consists of 14 items, and three subscales can be scored: ‘functional HL’ (five items), ‘communicative HL’ (five items), and ‘critical HL’ (four items) (Box S2). Responses to each item were scored on a four-point Likert scale, with scores ranging from 1 (never) to 4 (often). The scores were reversed to assess functional HL. For each of the three subscales, the average score of the corresponding item was used as the scale score. Scores ranged from 1 to 4, with higher scores indicating higher levels of HL. The scale demonstrated construct validity and reliability with high internal consistency for functional, communicative, and critical HL (coefficient alpha: 0.84, 0.77, and 0.65, respectively) in patients with type 2 diabetes in Japan.^19^

#### Trust in physicians

Trust in physicians was treated as a mediator (i.e., descendant of HL and ancestor of MA) and was measured using the five-item Japanese version of the Wake Forest Trust in Doctors Generally Scale.^20,21^ The scale consists of five items scored on a five-point Likert scale. The patients were asked to choose one answer for each item, ranging from ‘strongly disagree’ (1 point) to ‘strongly agree’ (5 points). After inverting the score for each negatively worded item, the sum of the scores was converted into a scale ranging from 0 to 100. The construct validity and internal consistency of the scale were verified with an alpha coefficient of 0.88.^20^

### Measurement of covariates

Confounding variables were selected based on the literature and expert medical knowledge and included those that were suspected to affect HL, trust in physicians, and MA. The variables included age, sex, smoking history, final education, household income, comorbid conditions (hypertension, diabetes, cardiovascular disease, liver disease, depression, and dementia), total number of classes of antihypertensive medications, and number of phosphate binders prescribed. Cardiovascular disease was defined as a history of myocardial infarction, angina pectoris, heart failure, cerebrovascular disease, or peripheral vascular disease. Hypertension was defined as taking antihypertensive medications regularly or having a systolic blood pressure of 140 mmHg or higher at the first dialysis session of each week (typically before starting dialysis on Monday or Saturday). A prescription of antihypertensives was considered present if any of the following was prescribed, where each antihypertensive agent was categorized into five classes for analysis: (i) calcium channel blockers, (ii) angiotensin-converting enzyme inhibitors, angiotensin II receptor blockers, angiotensin receptor neprilysin inhibitors, (L) alpha-beta blockers, beta blockers, (L) alpha blockers, and (L) central sympatholytic agents. The pill count for the phosphate binders was calculated if any of the following were prescribed: calcium-based, sevelamer, bixalomer, lanthanum, sucroferric oxyhydroxide, or ferrous citrate hydrate. The questionnaire was administered at each facility and patients were asked to complete the questionnaire at home.

### Statistical analysis

All statistical analyses were performed using Stata/SE, version 17 (StataCorp, College Station, TX, USA). Patient characteristics are described as median and interquartile range (IQR) for continuous variables and frequency and percentage for categorical variables. A series of general linear models were fitted to analyze the association of the trust in physicians with multidimensional HLs (see Figure 1 analysis #1), and those of the ASK-12 scores with HLs with/without trust in physicians (see Figure 1 analysis #2). Next, a series of mediation analyses was employed to determine whether trust in physicians mediates the relationship between multidimensional HLs and MA using Stata’s sgmediation2 command.^22^ The mediation analysis allowed us to estimate the effect of the independent variables (i.e., multidimensional HLs, i.e., the direct effect) after excluding the effect of the mediator (i.e., trust in the physician), the effect explained by the mediator (i.e., indirect effect), and the proportion mediated by the mediator. An outline of the mediation analyses is presented in Figure 2 and the details are described in Item S1.

**Figure 1.**
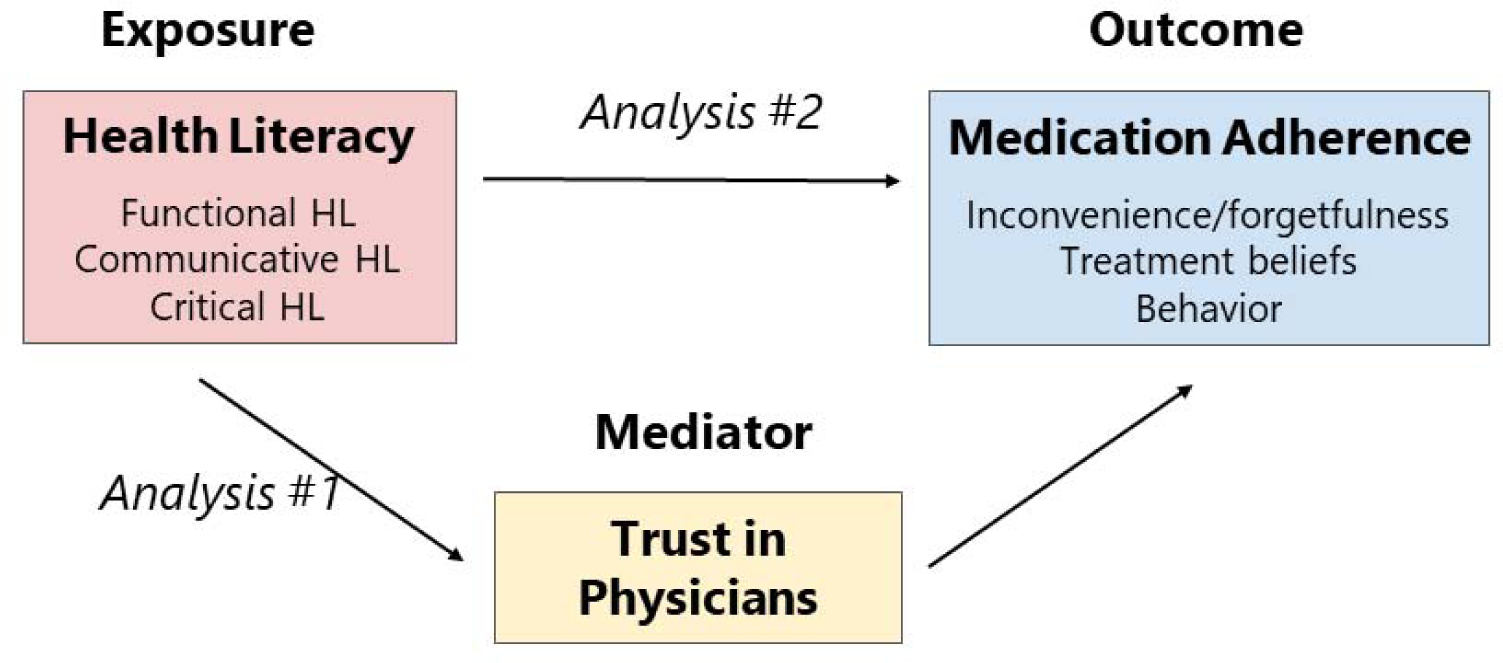
Conceptual framework for this study. Analysis #1: Association of trust in physicians and health literacy (HL). Analysis #2: Association of HL with medication adherence scores modeled with/without trust in physicians.

**Figure 2.**
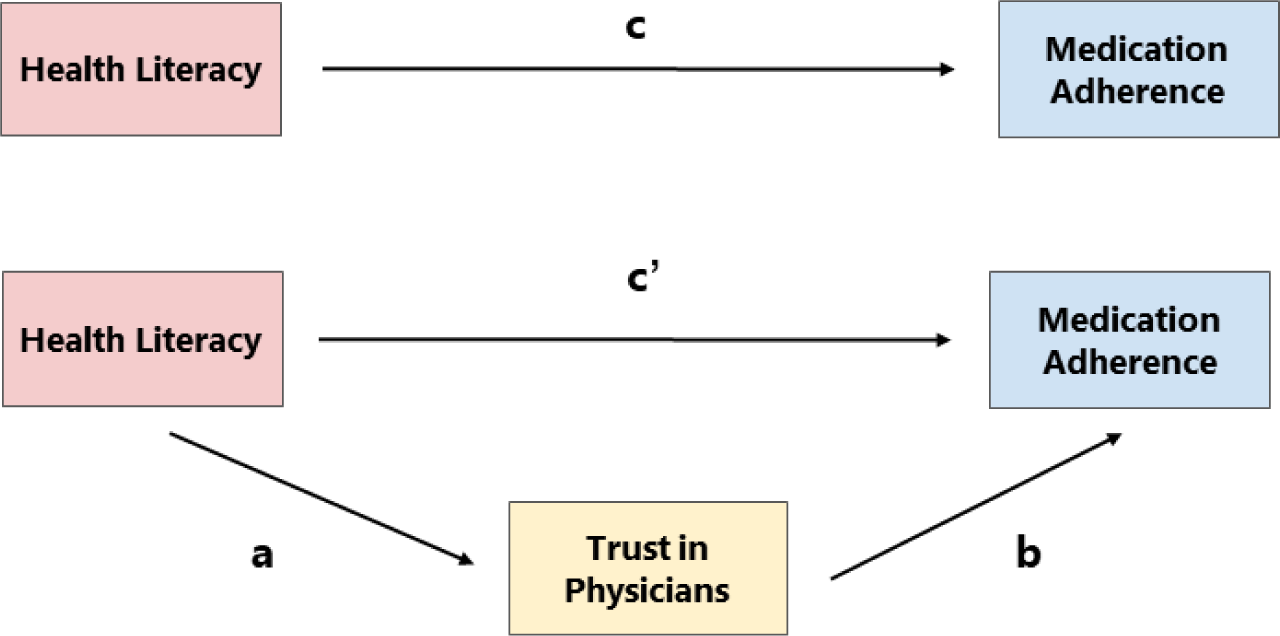
Study mediation analysis. First, medication adherence was regressed on health literacy (HL) and its covariates (excluding trust in physicians). The coefficient on the HL is c, and represents the “total effect” on medication adherence (i.e., the effect before removing the portion of the effect explained by trust in physicians) **(upper half of the figure)**. Second, the mediator, trust in physicians, was regressed on HL and the covariates. The coefficient of HL is P a. Third, medication adherence is regressed on HL, trust in physicians, and covariates. The coefficient of trust in physicians was b and the coefficient of HL was c’. The direct effect of the HL on medication adherence is denoted by c’. The indirect effect of HL on medication adherence through trust in physicians is represented by a × b. Therefore, c = c’ + a × b (**bottom half of the figure).**

The Sobel-Goodman test was used to determine whether trust in physicians significantly mediates the relationship between HL and MA. A relationship between HL and MA was considered completely mediated if the relationship was no longer significant after controlling for trust in physicians, and partially mediated if the relationship remained significant.^23^ This series of separate mediation analyses was performed for each association between functional, communicative, and critical HL and MA. In addition, the total score and each of the three sub-domain scores of MA were used for regression modeling. Ten imputations were performed using multiple imputations with chained equations, assuming that the analyzed data were randomly missing. Statistical significance was set at P < 0.05 all analyses.

## Results

### Study flow and participant characteristics

Of 651 patients undergoing hemodialysis at the seven centers, 484 (74.3%) participated in the study. Finally, 455 patients were included in the analysis after excluding those with missing ASK-12 responses (n=28) and one patient not regularly prescribed oral medications (n=1) (Figure 3). The participant characteristics are shown in Table 1. The median age was 71 years (IQR: 62–78 years) and 300 patients (65.9%) were male. The most common dialysis vintage was 5–9 years (29.0%). More than half (56.6%) of the patients were prescribed four or more phosphate binders. Two types of antihypertensive medications were most commonly prescribed (37.6%).

**Figure 3.**
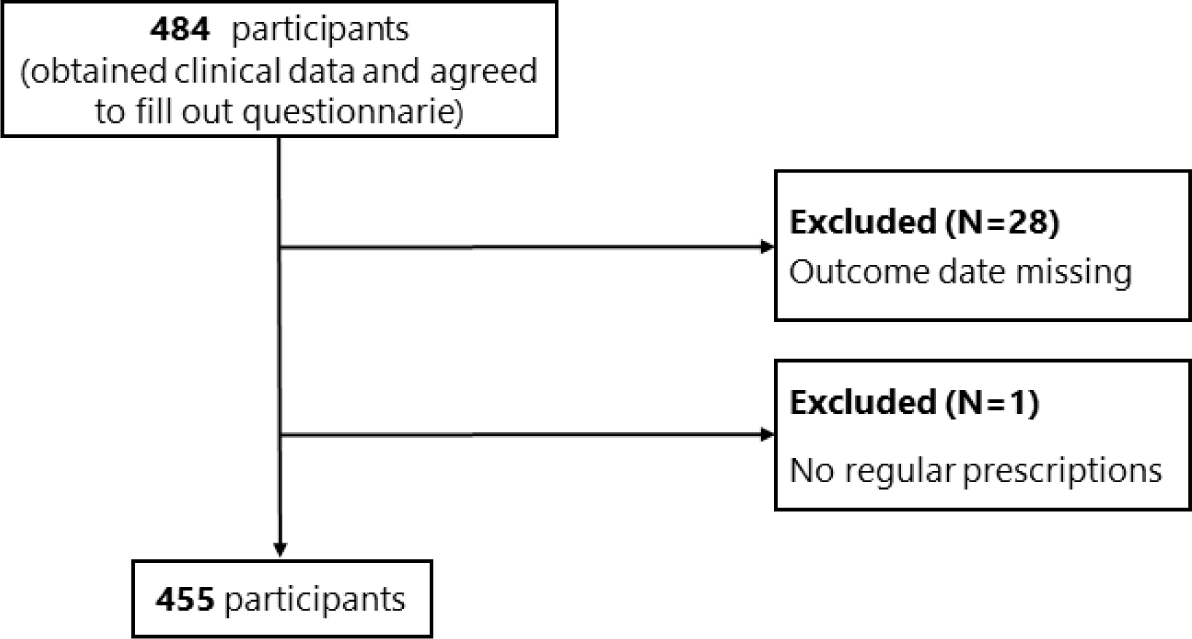
Study flow chart.

**Table 1.** Patient characteristics (n=455)

|  | **Total n=455** | Missing,<br>n |
| --- | --- | --- |
| Age (years) | 71 [62-78] | 0 |
| Men, n (%) | 300 (65.9) | 0 |
| Dialysis vintage, n (%) |  | 0 |
| *Less than 3 years* | 115 (25.3) |  |
| *3 to 4 years* | 86 (18.9) |  |
| *5 to 9 years* | 132 (29.0) |  |
| *10 to 14 years* | 59 (13.0) |  |
| *15 or more years* | 63 (13.8) |  |
| Renal disease, n (%) |  | 0 |
| *Diabetic nephropathy* | 171 (37.6) |  |
| *Chronic glomerulonephritis* | 106 (23.3) |  |
| *Nephrosclerosis* | 60 (13.2) |  |
| *Polycystic kidney disease* | 13 (2.9) |  |
| *Other* | 69 (15.1) |  |
| *Unknown* | 36 (7.9) |  |
| Comorbidities, n (%) |  |  |
| *Hypertension* | 349 (76.7) | 0 |
| *Diabetes* | 195 (43.1) | 3 |
| *Liver disease* | 27 (6.0) | 1 |
| *Cardiovascular disease* | 195 (43.2) | 4 |
| *Depression* | 10 (2.2) | 1 |
| *Dementia* | 20 (4.4) | 3 |
| Smoking, n (%) |  | 19 |
| *Never* | 233 (53.4) |  |
| *Ever* | 153 (35.1) |  |
| *Current* | 50 (11.5) |  |
| Education, n (%) |  | 20 |
| *Junior high school graduate or less* | 105 (24.2) |  |
| *High school graduate* | 212 (48.7) |  |
| *University/Graduate school graduate* | 61 (14.0) |  |
| *Others* | 57 (13.1) |  |
| Household income, n (%) |  | 33 |
| <1,000,000 yen | 41 (9.7) |  |
| 1,000,000 to <3,000,000 yen | 186 (44.1) |  |
| 3,000,000 to <5,000,000 yen | 111 (26.3) |  |
| 5,000,000 to <10,000,000 yen | 70 (16.6) |  |
| ≥10,000,000 yen | 14 (3.3) |  |
| Number of phosphorus binder, n (%) |  | 1 |
| *None* | 62 (13.7) |  |
| *1 to 3 tablets* | 135 (29.7) |  |
| *4 or more tablets* | 257 (56.6) |  |
| Number of antihypertensive classes, n (%) |  | 0 |
| *None* | 71 (15.6) |  |
| *One* | 126 (27.7) |  |
| *Two* | 171 (37.6) |  |
| *Three or more* | 87 (19.1) |  |

Continuous variables are summarized as median [interquartile range].
Categorical variables are summarized as frequencies and proportions (in parentheses).

### Association between HL and trust in physicians

The median trust in physicians score was 65 (IQR: 55–75), and the median of the functional HL, communicative HL, and critical HL subdomains were 3.2 (IQR: 2.6–3.8), 2.8 (IQR 2.2–3.2), and 2.5 (IQR 2.0–3.0), respectively. Table 2 shows the associations between trust in physicians in the HL subdomains and patient characteristics. Trust in physicians was positively associated with functional HL (per 1-point increase: 2.26 (95% CI: 0.04, 4.48)), communicative HL (per 1-point increase: 6.79 (95% CI: 3.49, 10.09)), and age (per 10-year increase: 2.48 (95% CI: 1.16, 3.81)). Critical HL (per 1-point increase: -7.76 (95% CI: -11.02, -4.51)) and education (especially college or graduate school) (reference to junior high school graduate or less: -6.22 (95% CI: -11.94, -0.50)) were inversely correlated with trust in physicians.

**Table 2.**
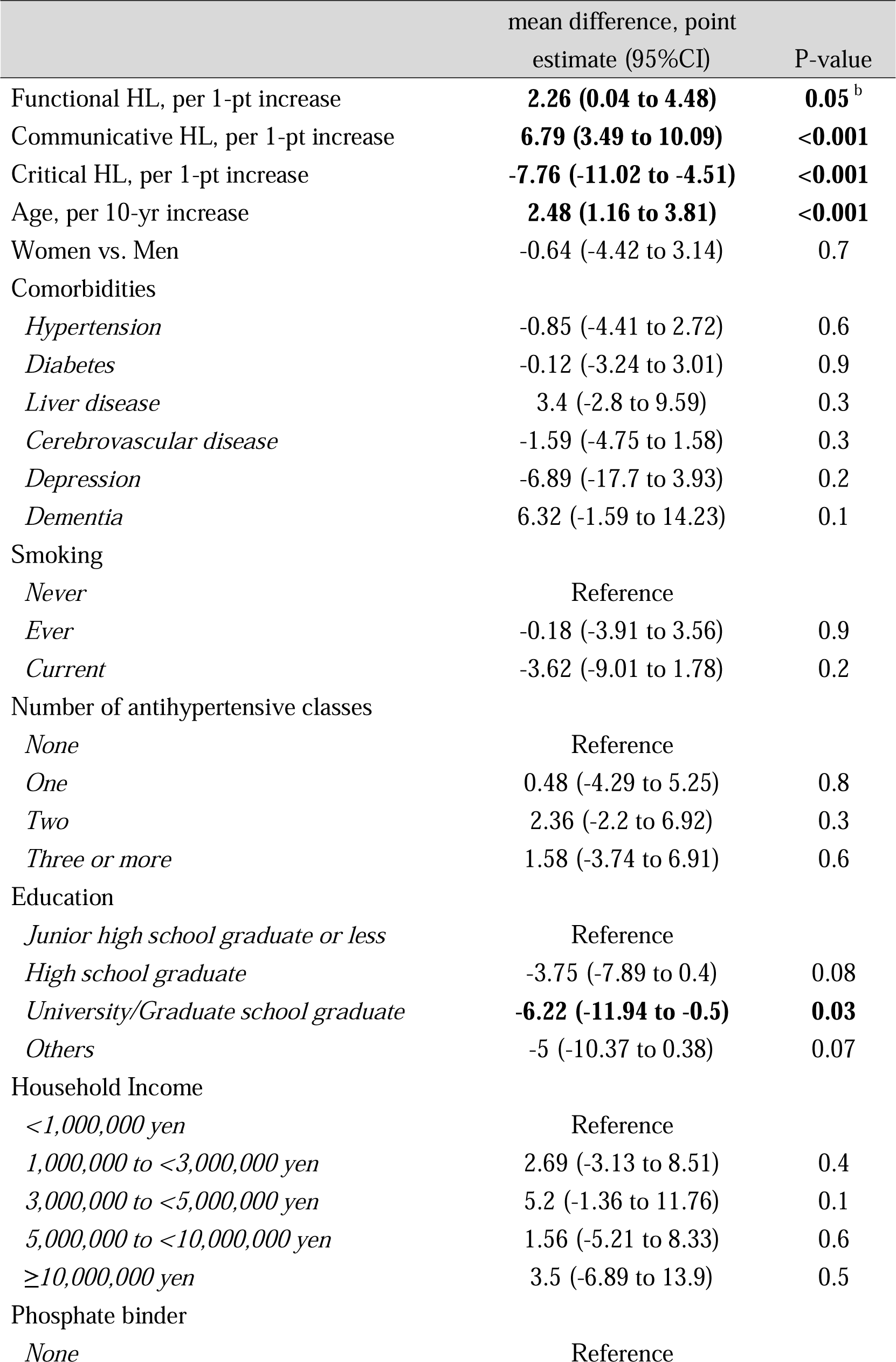
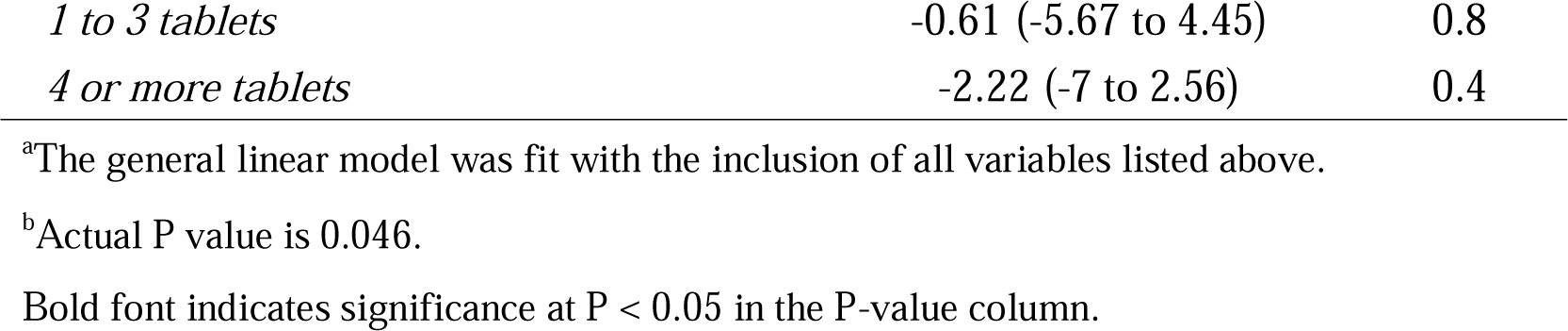
Association of trust in physicians with health literacy and other covariates^a^.

|  | mean difference, point<br>estimate (95%CI) | P-value |
| --- | --- | --- |
| Functional HL, per 1-pt increase | **2.26 (0.04 to 4.48)** | **0.05<sup>b</sup>** |
| Communicative HL, per 1-pt increase | **6.79 (3.49 to 10.09)** | **<0.001** |
| Critical HL, per 1-pt increase | **-7.76 (-11.02 to -4.51)** | **<0.001** |
| Age, per 10-yr increase | **2.48 (1.16 to 3.81)** | **<0.001** |
| Women vs. Men | -0.64 (-4.42 to 3.14) | 0.7 |
| Comorbidities |  |  |
| *Hypertension* | -0.85 (-4.41 to 2.72) | 0.6 |
| *Diabetes* | -0.12 (-3.24 to 3.01) | 0.9 |
| *Liver disease* | 3.4 (-2.8 to 9.59) | 0.3 |
| *Cerebrovascular disease* | -1.59 (-4.75 to 1.58) | 0.3 |
| *Depression* | -6.89 (-17.7 to 3.93) | 0.2 |
| *Dementia* | 6.32 (-1.59 to 14.23) | 0.1 |
| Smoking |  |  |
| *Never* | Reference |  |
| *Ever* | -0.18 (-3.91 to 3.56) | 0.9 |
| *Current* | -3.62 (-9.01 to 1.78) | 0.2 |
| Number of antihypertensive classes |  |  |
| *None* | Reference |  |
| *One* | 0.48 (-4.29 to 5.25) | 0.8 |
| *Two* | 2.36 (-2.2 to 6.92) | 0.3 |
| *Three or more* | 1.58 (-3.74 to 6.91) | 0.6 |
| Education |  |  |
| *Junior high school graduate or less* | Reference |  |
| *High school graduate* | -3.75 (-7.89 to 0.4) | 0.08 |
| *University/Graduate school graduate* | **-6.22 (-11.94 to -0.5)** | **0.03** |
| *Others* | -5 (-10.37 to 0.38) | 0.07 |
| Household Income |  |  |
| <1,000,000 yen | Reference |  |
| 1,000,000 to <3,000,000 yen | 2.69 (-3.13 to 8.51) | 0.4 |
| 3,000,000 to <5,000,000 yen | 5.2 (-1.36 to 11.76) | 0.1 |
| 5,000,000 to <10,000,000 yen | 1.56 (-5.21 to 8.33) | 0.6 |
| ≥10,000,000 yen | 3.5 (-6.89 to 13.9) | 0.5 |
| Phosphate binder |  |  |
| *None* | Reference |  |
| *1 to 3 tablets* | -0.61 (-5.67 to 4.45) | 0.8 |
| *4 or more tablets* | -2.22 (-7 to 2.56) | 0.4 |

<sup>a</sup>The general linear model was fit with the inclusion of all variables listed above.
<sup>b</sup>Actual P value is 0.046.
Bold font indicates significance at $P < 0.05$ in the P-value column.

### Association of MA with HL and trust in physicians

The median ASK-12 total score was 23 (IQR: 19–28), and the median ASK-12 subdomains were inconvenience/forgetfulness 6 (IQR: 4–8), belief 10 (IQR: 8–12), and behavior 6 (IQR: 5–9). Table 3 shows the associations between ASK-12 total score, multidimensional HLs, and trust in physicians. Higher functional and communicative HL were associated with lower barriers to MA (functional HL, per 1-point increase: -1.90 (95% CI: -2.67, -1.13), communicative HL, per 1-point increase: -2.11 (95% CI: -3.35, -0.87)); however, higher critical HL was associated with higher barriers (critical HL, per 1-point increase: 1.67 (95% CI: 0.43, 2.90); model without trust). Furthermore, when adjusted for trust in the physician, the strength of the association between communicative HL and MA became smaller (communicative HL, per 1-point increase: -1.32 (95% CI: -2.53, -0.10)), and the association disappeared completely for critical HL (critical HL, per 1-point increase: 0.76 (95% CI: -0.45, 1.98)). Table 4 shows the association between the HL and MA subdomains. Functional HL was positively associated with all MA domains. Higher communicative HL was positively associated with “belief”, and higher critical HL was negatively associated with “behavior” However, after adjusting for trust in physicians, the magnitude of the association between HL and MA decreased.

**Table 3.** Association of medication adherence score with health literacy and trust in physicians^a^ (n=455)

|  | Model WITHOUT trust<br>mean difference, point<br>estimate (95%CI) | P-value | Model WITH trust<br>mean difference, point<br>estimate (95%CI) | P-value |
| --- | --- | --- | --- | --- |
| Functional HL, per<br>1-pt increase | **-1.9 (-2.67 to -1.13)** | **<0.001** | **-1.64 (-2.38 to -0.89)** | **<0.001** |
| Communicative HL,<br>per 1-pt increase | **-2.11 (-3.35 to -0.87)** | **0.001** | **-1.32 (-2.53 to -0.1)** | **0.03** |
| Critical HL, per 1-pt<br>increase | **1.67 (0.43 to 2.9)** | **0.009** | 0.76 (-0.45 to 1.98) | 0.2 |
| Trust in physician,<br>per 10-pt increase |  |  | **-1.16 (-1.51 to -0.81)** | **<0.001** |

<sup>a</sup> The general linear model was fit with the inclusion of all variables listed in Table 2.
Bold font indicates significance at $P < 0.05$ in the P-value column.

**Table 4.** Association of medication adherence subdomains with health literacy and trust in physicians^a^ (n=455)

|  | Model WITHOUT trust |  | Model WITH trust |  |
| --- | --- | --- | --- | --- |
|  | mean difference, point estimate (95%CI) | P-value | mean difference, point estimate (95%CI) | P-value |
| **<u>Subdomain 1 = Inconvenience</u>** |  |  |  |  |
| Functional HL, per 1-pt increase | **-0.81 (-1.15 to -0.47)** | **<0.001** | **-0.75 (-1.09 to -0.41)** | **<0.001** |
| Communicative HL, per 1-pt increase | -0.3 (-0.86 to 0.26) | 0.3 | -0.13 (-0.69 to 0.44) | 0.7 |
| Critical HL, per 1-pt increase | 0.36 (-0.19 to 0.91) | 0.2 | 0.16 (-0.4 to 0.72) | 0.6 |
| Trust in physician, per 10-pt increase |  |  | **-0.26 (-0.41 to -0.1)** | **0.002** |
| **<u>Subdomain 2 = Belief</u>** |  |  |  |  |
| Functional HL, per 1-pt increase | **-0.51 (-0.87 to -0.15)** | **0.006** | **-0.39 (-0.73 to -0.04)** | **0.03** |
| Communicative HL, per 1-pt increase | **-1.34 (-1.92 to -0.77)** | **<0.001** | **-0.97 (-1.54 to -0.4)** | **0.001** |
| Critical HL, per 1-pt increase | 0.39 (-0.18 to 0.97) | 0.2 | -0.04 (-0.6 to 0.53) | 0.9 |
| Trust in physician, per 10-pt increase |  |  | **-0.55 (-0.73 to -0.38)** | **<0.001** |
| **<u>Subdomain 3 = Behavior</u>** |  |  |  |  |
| Functional HL, per 1-pt increase | **-0.58 (-1.03 to -0.13)** | **0.01** | **-0.5 (-0.95 to -0.06)** | **0.03** |
| Communicative HL, per 1-pt increase | -0.46 (-1.19 to 0.27) | 0.2 | -0.22 (-0.96 to 0.51) | 0.6 |
| Critical HL, per 1-pt increase | **0.91 (0.18 to 1.64)** | **0.01** | 0.64 (-0.1 to 1.37) | 0.09 |
| Trust in physician, per 10-pt increase |  |  | **-0.35 (-0.57 to -0.13)** | **0.002** |

<sup>a</sup> The general linear model was fit with the inclusion of all variables listed in Table 2.
Bold font indicates significance at $P < 0.05$ in the P-value column.

### Mediation effects of trust in physicians on the relationships between HL and MA

Table 5 and Figure S1 present the results of the mediation analyses. Functional HL was positively associated with total score and “belief” in MA, and trust in physicians was partially mediated by 15.4% and 28.4% of each association, respectively. Similarly, communicative HL was positively associated with total score and “belief” in MA, and trust in physicians was partially mediated by 35.0% and 21.6% of each association, respectively. Critical HL was inversely associated with total score and “behavior” in MA, and trust in physicians was completely mediated by 54.0% and 30.4% of each association, respectively.

**Table 5.** Summary of mediating effects of health literacy on medication adherence.

|  | Health literacy (HL) domain |  |  |
| --- | --- | --- | --- |
|  | Functional HL | Communicative HL | Critical HL |
| **<u>Total score</u>** |  |  |  |
| Direct effect (c') | -1.512 | -1.535 | 0.784 |
| Indirect effect (ab) | -0.275 | -0.828 | 0.921 |
| Total effect (c) | -1.786 | -2.363 | 1.705 |
| Percent of indirect effect | 15.4 % | 35 % | 54 % |
| Mediation | Partially mediated | Partially mediated | Completely mediated |
| Sobel test | p = 0.04 | p < 0.001 | p < 0.001 |
| **<u>Subdomain 1 = Inconvenience</u>** |  |  |  |
| Direct effect (c') | -0.709 |  |  |
| Indirect effect (ab) | -0.061 |  |  |
| Total effect (c) | -0.77 | Not associated | Not associated |
| Percent of indirect effect | 8 % |  |  |
| Mediation | Not mediated | N/A | N/A |
| Sobel test | p = 0.08 |  |  |
| **<u>Subdomain 2 = Belief</u>** |  |  |  |
| Direct effect (c') | -0.327 | -1.001 |  |
| Indirect effect (ab) | -0.13 | -0.391 |  |
| Total effect (c) | -0.456 | -1.393 | Not associated |
| Percent of indirect effect | 28.4 % | 28.1 % |  |
| Mediation | Partially mediated | Partially mediated | N/A |
| Sobel test | p = 0.04 | p < 0.001 |  |
| **<u>Subdomain 3 = Behavior</u>** |  |  |  |
| Direct effect (c') | -0.476 |  | 0.641 |
| Indirect effect (ab) | -0.084 |  | 0.28 |
| Total effect (c) | -0.56 | Not associated | 0.921 |
| Percent of indirect effect | 14.9 % |  | 30.4 % |
| Mediation | Not mediated | N/A | Completely mediated |
| Sobel test | p = 0.08 |  | p = 0.007 |

N/A; Not applicable.

## Discussion

This study determined how multidimensional HL is associated with MA among patients undergoing hemodialysis and tested whether trust in physicians mediates these associations. We found that high functional and communicative HL were associated with good MA, whereas high critical HL was associated with poor MA. Furthermore, communicative and critical HLs were positively and negatively associated with trust in physicians, respectively, and trust in physicians mediated this association. The present results underscore the need for interventions tailored to the patient’s HL level to improve MA and provide deep insights into potential interventions both within and outside of the physician–patient partnership formation.

The association between multidimensional HLs and MA in this study reinforces the findings of previous studies. A study on hemodialysis in Australia using the same multidimensional HLs as our study showed that each domain was positively associated with MA.^10^ However, the MA instrument used in the previous study had not been adequately validated. In addition, the participation rate in the study was <30%, and the mean age of the participants was 50 years. Therefore, the associations between multidimensional HL and MA demonstrated in that study have limited generalizability. In contrast, a study on CKD, including hemodialysis, from Norway found negative findings on the association between multidimensional HLs measured using the Health Literacy Questionnaire 47 and MA.^9^ However, the ceiling effect of the MA scale used in that study (median score of 24 out of a maximum score of 25) may have led to failure to detect differences in MA according to HLs.

The mediating role of trust in physicians on the association between multidimensional HL and MA has not yet been quantified. Our demonstration of the association between communicative HL and trust in physicians reinforces the hypothesis of the qualitative study, suggesting that increased daily communication between patients with CKD and their physicians lowers the patients’ perceived threshold for discussing their health concerns.^15^ Thus, higher communicative HL may help create more opportunities to receive explanations from physicians and foster feelings of security and trust in physicians. Moreover, the present finding of an inverse association between critical HL and trust in physicians indicates that individuals with higher critical HL may be more skeptical of medical care because they are suspicious of the reliability of the information, as indicated in the item. It also reinforces the hypothesis in the aforementioned qualitative study that patients with CKD trust the information provided by their physicians and do not continue thinking and worrying because of poor disease knowledge.^15^ A similar association has been observed in patients with SLE.^14^ In contrast, only one qualitative study on CKD suggests the importance of the role of trust in physicians in MA; suboptimal physician interactions with patients can compromise trust in physicians’ recommendations.^24^ As a result, patients hide their medication concerns, leading to nonadherence. Conversely, patients who developed a trusting relationship with their physicians adhered to the prescription instructions with little resistance.

These findings highlight the need for researchers and dialysis physicians to focus on differences in individual HL and the presence of physician mediation to devise individualized interventions to improve MA.

The partial mediating role of trust in physicians in the association between communicative HL and favorable MA (especially “beliefs”) indicates potential opportunities for improving MA through both physician–patient interaction and other means. Physicians should create a welcoming atmosphere and provide sufficient time for dialogue to attentively solicit patients’ medication preferences and concerns. The importance of physicians’ attitudes is supported by several qualitative studies suggesting that a lack of physicians’ assessment of adverse effects and empathy can lead to patients’ distrust of their physicians. First, inadequate physician–patient interactions lead to patients avoiding discussions with their physicians.

Consequently, patients’ “belief” in and adherence to medication are compromised as a result of confusion about these medications and unresolved fears.^24–26^ Additionally, because hemodialysis patients spend a lot of time interacting with nurses and other medical personnel in addition to their physicians, these professionals can foster a dialogue about their concerns and experiences with their medications, thereby providing an opportunity for patients to better understand medication-related information and improve their adherence. Second, the complete mediating role of trust in physicians in the association between critical HL and poor MA (especially “behavior”) indicates the sole importance of concentrating on a good physician–patient interaction for patients with high critical HL. Patients with higher critical HL may be more likely to complain and seek the opinions of other physicians.^27^ Therefore, physicians’ provision of solid knowledge about the medication (i.e., maintaining competence) and their attitude of aligning with the patient’s individuality and doing their best (i.e., ensuring reliability and dependability) may improve behaviors toward prescriptions based on suspicion and vigilance.^12^ Third, the partial mediating role of trust in physicians in the association between functional HL and good MA (especially “beliefs”) suggests the necessity for a concerted approach by both physicians and other healthcare professionals, as in the case of communicative HL. Considering items in functional HL, slow speech in plain language and visual aids to complement verbal explanations can facilitate medication comprehension.^28^ In fact, the latter has been shown to improve CKD patients’ knowledge and treatment decision-making.^29^ For elderly patients with hearing loss, physicians’ use of real-time text transcription of conversations using smartphone applications may facilitate patient comprehension and convey compassion to the patient, thereby helping build trust in the physician.

This study has several strengths. First, the multicenter design ensures the generalizability of our findings. Second, formal mediation analysis allowed us to quantify the extent to which trust in physicians mediated the association between multidimensional HLs and MA. Third, using the ASK-12, the association between the three detailed barriers to MA and multidimensional HLs could be analyzed. This enabled us to devise specific strategies according to the barriers to MA.

This study also has several limitations. First, owing to the cross-sectional nature of the study, the possibility of reverse causality cannot be excluded. Second, the MA assessment is based on self-reported instruments and does not measure pill counts or regimen complexity.^30,31^ However, we excluded individuals without regular prescriptions and adjusted our analysis for the pill counts of phosphate binders and number of classes of antihypertensive medications as proxies for pill counts. In contrast, an objective electronic monitoring system, such as a Medication Event Monitoring System (MEMS), can be used.^32^ However, the use of MEMS was not feasible in this population-based study because of the large number of pills administered to hemodialysis patients and the high cost of the equipment. Third, the trust in physicians scale used in this study may not capture trust in one physician, as it specifically targets trust in physicians.^20^ However, in Japan, more than one physician visits hemodialysis patients on a shift basis, even at the same facility. Therefore, we did not use a scale to measure the trust in a single attending physician.

In conclusion, among patients undergoing hemodialysis, higher communicative HL was associated with better MA, whereas higher critical HL was associated with worse MA, independent of functional HL. Furthermore, trust in physicians mediated the associations between all three HLs and MA and, in particular, completely mediated the association between critical HL and MA. To effectively improve MA, it is important to establish interventions tailored to individual HL levels and favorable physician–patient partnerships.

## Supplementary Material

Box S1. Japanese version of the Adherence Starts with Knowledge 12 Scale (ASK-12)

Box S2. The Functional Communicative Critical Health Literacy Scale (FCCHL)

Item S1. Brief description of the mediation analysis

Figure S1. Summary of the degree to which trust in physicians mediates the association between health literacy and medication adherence total score

## Article Information

### Authors’ Contributions

Research idea and study design: RI, T. Toida, NK; data acquisition: RI, YK, MU, T. Toishi, AK, MM, TO, YM, TS; data analysis/interpretation: TA, NK; statistical analysis: RI, T. Toida, NK; supervision or mentorship: T. Toida, NK. Each author contributed important intellectual content during manuscript drafting or revision, agreed to be personally accountable for the individual’s own contributions, and ensured that questions pertaining to the accuracy or integrity of any portion of the work, even one in which the author was not directly involved, were appropriately investigated and resolved, including documentation in the literature, if appropriate.

### Support

This study was supported by JSPS KAKENHI (grant numbers: JP19KT0021 and JP23K16271).

## Data Availability

All data produced in the present work are contained in the manuscript.

## Acknowledgements

The authors greatly thank the following researchers, research assistants, and medical staff members for their assistance in collecting the questionnaire-based and clinical information used in this study: Ms. Aki Tairaku (Shin-Yurigaoka General Hospital, Kawasaki-City, Kanagawa); Ms. Takako Saruwatari and Ms. Akiko Kamimura (Kyushu University of Health and Welfare, Nobeoka-City, Miyazaki); Tetuo Ueki, MD, Akio Munakata, MD, Yoshihiko Watanabe, MD (Munakata Clinic, Mobara-City, Chiba); Ms. Yayoi Takanashi, Reiji Masaki, NP, Tomohiko Inoue, MD, Shinnosuke Sugihara, MD, Kanako Nagaoka, MD and Hiroshi Kuji, MD (Kameda Medical Center, Kamogawa-City, Chiba); Kenji Yamaguchi, MD (Awa Regional Medical Center, Tateyama-City, Chiba); Ms. Miyuki Sato (Fukushima Medical University Hospital, Fukushima-City, Fukushima).

## References

1. Mason NA, Bakus JL. Strategies for reducing polypharmacy and other medication-related problems in chronic kidney disease. Semin Dial. 2010;23(1):55–61. doi:10.1111/j.1525-139X.2009.00629.x

2. Manley HJ, Garvin CG, Drayer DK, et al. Medication prescribing patterns in ambulatory haemodialysis patients: comparisons of USRDS to a large not-for-profit dialysis provider. Nephrol Dial Transplant. 2004;19(7):1842–1848. doi:10.1093/ndt/gfh280

3. Chiu YW, Teitelbaum I, Misra M, de Leon EM, Adzize T, Mehrotra R. Pill burden, adherence, hyperphosphatemia, and quality of life in maintenance dialysis patients. Clin J Am Soc Nephrol. 2009;4(6):1089–1096. doi:10.2215/CJN.00290109

4. Schmid H, Hartmann B, Schiffl H. Adherence to prescribed oral medication in adult patients undergoing chronic hemodialysis: a critical review of the literature. Eur J Med Res. 2009;14(5):185–190. doi:10.1186/2047-783x-14-5-185

5. Molnar MZ, Gosmanova EO, Sumida K, et al. Predialysis Cardiovascular Disease Medication Adherence and Mortality After Transition to Dialysis. Am J Kidney Dis. 2016;68(4):609–618. doi:10.1053/j.ajkd.2016.02.051

6. Fissell RB, Karaboyas A, Bieber BA, et al. Phosphate binder pill burden, patient-reported non-adherence, and mineral bone disorder markers: Findings from the DOPPS. Hemodial Int. 2016;20(1):38–49. doi:10.1111/hdi.12315

7. Fraser SDS, Roderick PJ, Casey M, Taal MW, Yuen HM, Nutbeam D. Prevalence and associations of limited health literacy in chronic kidney disease: a systematic review. Nephrol Dial Transplant. 2013;28(1):129–137. doi:10.1093/ndt/gfs371

8. Nutbeam D, McGill B, Premkumar P. Improving health literacy in community populations: a review of progress. Health Promot Int. 2018;33(5):901–911. doi:10.1093/heapro/dax015

9. Stømer UE, Wahl AK, Gøransson LG, Urstad KH. Health Literacy in Kidney Disease: Associations with Quality of Life and Adherence. J Ren Care. 2020;46(2):85–94. doi:10.1111/jorc.12314

10. Indino K, Sharp R, Esterman A. The effect of health literacy on treatment adherence in maintenance haemodialysis patients: a cross-sectional study. Ren Soc Australas J. 2019;15(1):11–18. doi:10.33235/rsaj.15.1.11-18

11. Paasche-Orlow MK, Wolf MS. The causal pathways linking health literacy to health outcomes. Am J Health Behav. 2007;31 Suppl 1:S19–S26. doi:10.5555/ajhb.2007.31.supp.S19

12. Pearson SD, Raeke LH. Patients’ trust in physicians: many theories, few measures, and little data. J Gen Intern Med. 2000;15(7):509–513. doi:10.1046/j.1525-1497.2000.11002.x

13. Kurita N, Oguro N, Miyawaki Y, et al. Trust in the attending rheumatologist, health-related hope and medication adherence among Japanese systemic lupus erythematosus patients. Rheumatology (Oxford). 2023;62(6):2147–2153. doi:10.1093/rheumatology/keac565

14. Oguro N, Yajima N, Miyawaki Y, et al. Effect of Communicative and Critical Health Literacy on Trust in Physicians Among Patients With Systemic Lupus Erythematosus (SLE): The TRUMP2-SLE Project. J Rheumatol. 2023;50(5):649–655. doi:10.3899/jrheum.220678

15. Stømer UE, Wahl AK, Gøransson LG, Urstad KH. Exploring health literacy in patients with chronic kidney disease: a qualitative study. BMC Nephrol. 2020;21(1):314. doi:10.1186/s12882-020-01973-9

16. Matza LS, Park J, Coyne KS, Skinner EP, Malley KG, Wolever RQ. Derivation and Validation of the ASK-12 Adherence Barrier Survey. Ann Pharmacother. 2009;43(10):1621–1630. doi:10.1345/aph.1M174

17. Takemura M, Nishio M, Fukumitsu K, et al. Optimal cut-off value and clinical usefulness of the Adherence Starts with Knowledge-12 in patients with asthma taking inhaled corticosteroids. J Thorac Dis. 2017;9(8):2350–2359. doi:10.21037/jtd.2017.06.115

18. Ito H, Hiramatsu T, Kawai K. Association between adherence and treatment satisfaction in adult patients with asthma. J Jp Soc Resp Care Rehab. 2019;28(1):97–102 (In Japanese). doi:10.15032/jsrcr.28.1_97

19. Ishikawa H, Takeuchi T, Yano E. Measuring functional, communicative, and critical health literacy among diabetic patients. Diabetes Care. 2008;31(5):874–879. doi:10.2337/dc07-1932

20. Oguro N, Suzuki R, Yajima N, et al. The impact that family members’ health care experiences have on patients’ trust in physicians. BMC Health Serv Res. 2021;21(1):1122. doi:10.1186/s12913-021-07172-y

21. Dugan E, Trachtenberg F, Hall MA. Development of abbreviated measures to assess patient trust in a physician, a health insurer, and the medical profession. BMC Health Serv Res. 2005;5:64. doi:10.1186/1472-6963-5-64

22. Mize TD. sgmediation2 - Sobel-Goodman Tests of Mediation in Stata. Accessed August 8, 2023. https://www.trentonmize.com/software/sgmediation2

23. MacKinnon DP, Fairchild AJ, Fritz MS. Mediation analysis. Annu Rev Psychol. 2007;58:593-614. doi:10.1146/annurev.psych.58.110405.085542

24. Ghimire S, Castelino RL, Jose MD, Zaidi STR. Medication adherence perspectives in haemodialysis patients: a qualitative study. BMC Nephrol. 2017;18(1):167. doi:10.1186/s12882-017-0583-9

25. Mechta Nielsen T, Frøjk Juhl M, Feldt-Rasmussen B, Thomsen T. Adherence to medication in patients with chronic kidney disease: a systematic review of qualitative research. Clin Kidney J. 2018;11(4):513–527. doi:10.1093/ckj/sfx140

26. Rifkin DE, Laws MB, Rao M, Balakrishnan VS, Sarnak MJ, Wilson IB. Medication adherence behavior and priorities among older adults with CKD: a semistructured interview study. Am J Kidney Dis. 2010;56(3):439–446. doi:10.1053/j.ajkd.2010.04.021

27. Bertram M, Brandt US, Hansen RK, Svendsen GT. Does higher health literacy lead to higher trust in public hospitals? Int J Equity Health. 2021;20(1):209. doi:10.1186/s12939-021-01528-w

28. Weiss BD. Health Literacy and Patient Safety: Help Patients Understand. Manual for Clinicians. Second edition. American Medical Association Foundation; 2007.

29. Patzer RE, McPherson L, Basu M, et al. Effect of the iChoose Kidney decision aid in improving knowledge about treatment options among transplant candidates: A randomized controlled trial. Am J Transplant. 2018;18(8):1954–1965. doi:10.1111/ajt.14693

30. Cardone KE, Manley HJ, Grabe DW, Meola S, Hoy CD, Bailie GR. Quantifying home medication regimen changes and quality of life in patients receiving nocturnal home hemodialysis. Hemodial Int. 2011;15(2):234–242. doi:10.1111/j.1542-4758.2011.00539.x

31. Neri L, Martini A, Andreucci VE, et al. Regimen complexity and prescription adherence in dialysis patients. Am J Nephrol. 2011;34(1):71–76. doi:10.1159/000328391

32. Nguyen TMU, Caze AL, Cottrell N. What are validated self-report adherence scales really measuring?: a systematic review. Br J Clin Pharmacol. 2014;77(3):427–445. doi:10.1111/bcp.12194

