## Supplementary Files for "The mediating role of trust in physicians on the association between multidimensional health literacy and medication adherence in hemodialysis: A cross-sectional study"

**Supplementary Box S1. The Japanese version of the Adherence Starts with Knowledge 12 scale (ASK-12)** ^1,2^

| Instruction sentence | English: “Taking Medicine-What Gets in the Way? Think about all of the medicines you take. Mark one answer for each item below.” |
| --- | --- |
| English: “Lifestyles” | |
| Question 1 | English: “I forget to take my medicines some of the time.” |
| Question 2 | English: “I run out of my medicines because I don’t get refills on time.” |
| Question 3 | English: “Taking medicines more than once a day is inconvenient.” |
| English: “Attitudes and Beliefs” | |
| Question 4 | English: “I feel confident that each one of my medicines will help  me.” |
| Question 5 | English: “I know if I am reaching my health goals.” |
| English: “Help From Others” | |
| Question 6 | English: “I have someone whom I can call with questions about  my medicines.” |
| English: “Talking With Healthcare Team” | |
| Question 7 | English: “My doctor/nurse and I work together to make decisions.” |
| English: “Taking Medicines” | |
| Question 8 | English: “Have you taken a medicine more or less often than  prescribed?” |
| Question 9 | English: “Have you skipped or stopped taking a medicine  because you didn’t think it was working?” |
| Question 10 | English: “Have you skipped or stopped taking a medicine  because it made you feel bad?” |
| Question 11 | English: “Have you skipped, stopped, not refilled, or taken less  medicine because of the cost?” |
| Question 12 | English: “Have you not had medicine with you when it was time  to take it?” |

The English-translated version ^3^ is also provided for each item and response.

Questions 1 through 3 constitute the “Inconvenience/Forgetfulness” domain, questions 4 through 7 the “Treatment Beliefs” domain, and questions 8 through 12 the “Behavior” domain.

Due to the medRxiv policy non-English information cannot be posted, thus please access this link for information on the items for the Japanese version: https://www.jstage.jst.go.jp/article/jsrcr/28/1/28_97/_pdf/-char/ja

Reference

1.	Takemura M, Nishio M, Fukumitsu K, et al. Optimal cut-off value and clinical usefulness of the Adherence Starts with Knowledge-12 in patients with asthma taking inhaled corticosteroids. *J Thorac Dis*. 2017;9(8):2350-2359. doi:[10.21037/jtd.2017.06.115](http://dx.doi.org/10.21037/jtd.2017.06.115)

2.	Ito H, Hiramatsu T, Kawai K. Association between adherence and treatment satisfaction in adult patients with asthma. *J Jp Soc Resp Care Rehab*. 2019;28(1):97-102 (In Japanese). doi:[10.15032/jsrcr.28.1_97](http://dx.doi.org/10.15032/jsrcr.28.1_97)

3.	Matza LS, Park J, Coyne KS, Skinner EP, Malley KG, Wolever RQ. Derivation and Validation of the ASK-12 Adherence Barrier Survey. *Ann Pharmacother*. 2009;43(10):1621-1630. doi:[10.1345/aph.1M174](http://dx.doi.org/10.1345/aph.1M174)

**Supplementary Box S2. The Functional Communicative Critical Health Literacy Scale (FCCHL)**

| Instruction sentence | English: “In reading instructions or leaflets from hospitals/pharmacies, you. . .” |
| --- | --- |
| Question 1 | English: “found that the print was too small to read.” |
| Question 2 | English: “found characters and words that you did not know” |
| Question 3 | English: “found that the content was too difficult.” |
| Question 4 | English: “needed a long time to read and understand them.” |
| Question 5 | English: “needed someone to help you read them.” |
| Instruction sentence | English: “Since having systemic lupus erythematosus you have. . .” |
| Question 1 | English: “collected information from various sources.” |
| Question 2 | English: “extracted the information you wanted.” |
| Question 3 | English: “understood the obtained information.” |
| Question 4 | English: “communicated your thoughts about your illness to someone.” |
| Question 5 | English: “applied the obtained information to your daily life.” |
| Question 6 | English: “considered whether the information was applicable to your situation.” |
| Question 7 | English: “considered the credibility of the information.” |
| Question 8 | English: “checked whether the information was valid and reliable.” |
| Question 9 | English: “collected information to make health-related decisions.” |
| Response options for questions | English: “never / rarely / sometimes / often” |

The English-translated version [1] is provided for each item and response. Due to the medRxiv policy, non-English information cannot be posted, thus please access this link to obtain the Japanese version: https://www.medrxiv.org/content/medrxiv/early/2022/05/16/2022.05.13.22275070/DC1/embed/media-1.pdf?download=true

**Reference**

1. Ishikawa H, Takeuchi T, Yano E (2008) Measuring Functional, Communicative, and Critical Health Literacy Among Diabetic Patients. *Diabetes Care* 31 (5):874-879. doi:[10.2337/dc07-1932](https://doi.org/10.2337/dc07-1932).

**Item S1: Brief description of mediation analysis**

Using the aforementioned general linear models, we then estimated and tested the paths discussed in the following ([see](https://docs.google.com/presentation/d/1lUugTom280vvaavrXMS0fREdGr7Vxly0y_SAys3LACc/edit#slide=id.p) [Figure2](https://docs.google.com/presentation/d/1lUugTom280vvaavrXMS0fREdGr7Vxly0y_SAys3LACc/edit#slide=id.g25106dd9ddc_0_0)). First, medication adherence is regressed on the HL and the covariates (other than trust in physicians). The coefficient on the HL is c, and represents the "total effect" on medication adherence (i.e., the effect before removing the portion of the effect explained by trust in physicians). Second, the mediator - trust in physicians - is regressed on the HL and the covariates. The coefficient on the HL is path a. Third, medication adherence is regressed on the HL, trust in physicians, and the covariates. The coefficient on trust in physicians is b, and the coefficient on the HL is c'. The direct effect of the HL on medication adherence is denoted by c'. The indirect effect of the HL on medication adherence through trust in physicians is represented by a × b. The Sobel-Goodman test was performed to determine whether trust in physicians significantly mediates the relationship between the HL and MA. A relationship between the HL and MA was considered completely mediated if the relationship was no longer significant after controlling for trust in physicians, and partially mediated if the relationship remained significant.^1^ This series of separate mediation analysis was performed for each of the associations of functional HL, communicative HL, and critical HL with MA, respectively. In addition, a total score and each of the three sub-domain scores of MA were used for a regression modeling.

Reference

1.	MacKinnon DP, Fairchild AJ, Fritz MS. Mediation analysis. *Annu Rev Psychol*. 2007;58:593-614. doi:[10.1146/annurev.psych.58.110405.085542](http://dx.doi.org/10.1146/annurev.psych.58.110405.085542)

**Supplementary Figure 1. Summary of the degree to which trust in physicians mediates the association between health literacy and medication adherence total score.**

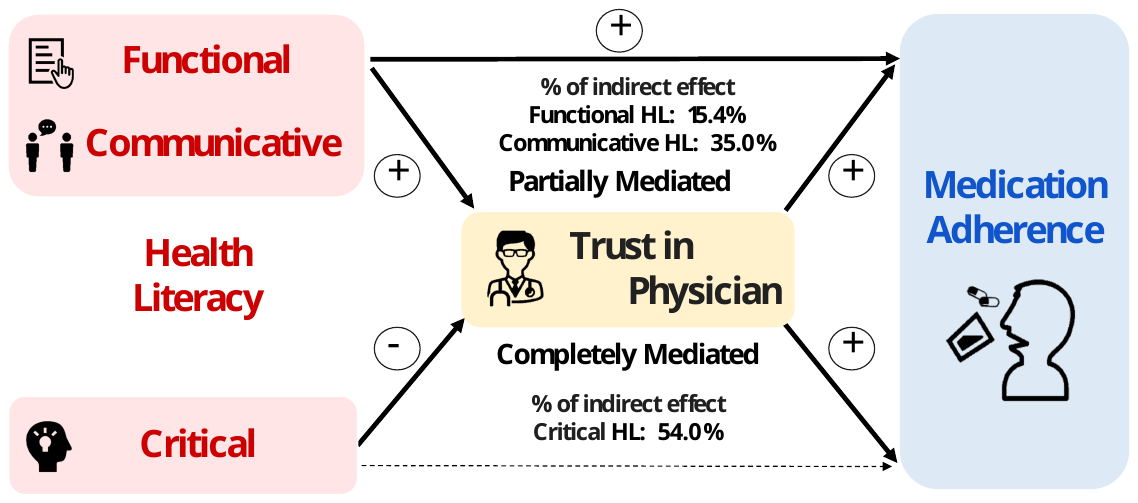
